# Indications for healthcare surge capacity in European countries facing an exponential increase in COVID19 cases

**DOI:** 10.1101/2020.03.14.20035980

**Authors:** Frederik Verelst, Elise Kuylen, Philippe Beutels

**Affiliations:** Centre for Health Economics Research & Modelling Infectious Diseases (CHERMID), Vaccine and Infectious Disease Institute (VAXINFECTIO), University of Antwerp, Antwerp, Belgium; Interuniversity Institute for Biostatistics and statistical Bioinformatics, Hasselt University, Belgium; School of Public Health and Community Medicine, The University of New South Wales, Sydney, Australia

**Keywords:** COVID-19, healthcare, surge capacity, Italy, ICU

## Abstract

European healthcare systems face extreme pressure from COVID-19. We estimate such pressure by relating both country-specific accumulated COVID-19 deaths (intensity-approach) and active COVID-19 cases (magnitude-approach) to measures of healthcare system capacity: hospital beds, healthcare workers and healthcare expenditure. On March 25, 2020 - relative to Italy on March 11- we found Spain, The Netherlands and France to experience the highest pressure using the intensity-approach with a composite measure for healthcare capacity. For updates see www.covid-hcpressure.org

---

In the past weeks, it has become clear the SARS-COV-2 poses an humongous health threat on a global scale. Europe is currently the most affected continent in terms of detected COVID-19 cases and deaths. The outbreak initially took off the fastest in Italy, but has since spread to all other EU countries. We aim to track pressure on healthcare systems in the EU by comparing healthcare base capacity to the impact of COVID-19 in terms of cumulative deaths over the past 21 days and reported active cases. The results of our analyses are updated in real-time at www.covid-hcpressure.org

## Flattening the curve to reduce healthcare pressure in the EU

Several institutions have communicated the necessity to ‘flatten the curve’ in order to lower pressure on health care institutions and to buy time for antivirals and other medication to become available in the short term and vaccines in the longer term. Remuzzi & Remuzzi reported on March 12th that in Italy ICU bed occupation would exceed capacity [1]. From around the same time, other countries increasingly implemented large scale interventions such as household isolation and school closure [2, 3]. Important urgent questions as the epidemic unfolds are (a) how close other countries are from reaching an Italy-like pressure on the health system, (b) which countries are closest to such a situation, and (c) what pressure will countries experience at the peak and how will that pressure affect treatment of cases. Furthermore, policy makers require information on the magnitude of additional health care capacity that is needed when at a later stage, new exponential growth may occur after some of the large scale interventions are gradually eased and, perhaps, re-implemented.

## Healthcare base capacity measures in Europe

We characterized country-level healthcare base capacity by four measures:

1. **Hospital beds**. Three categories of hospital beds were considered: (1) available hospital bed capacity, (2) curative hospital bed capacity and (3) critical care hospital bed capacity.
2. **Physicians**. (1) the number of physicians and (2) the number of generalist medical practitioners.
3. **Composite measures**. In order to represent both healthcare workers and critical care beds, we composed two composite measures:

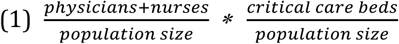

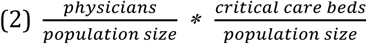
4. **Healthcare expenditures**. Data on the healthcare expenditure as a % of GDP.

Data on healthcare resources were derived from Eurostat [4] for the most recent year available for each country. We used data on the number of “Practicing nurses” where available. We retrieved data on “Professionally active nurses” for Ireland, France, Portugal and Slovakia. Critical care hospital bed capacity was extracted from a paper by Rhodes et al. [5].

## Intensity approach and magnitude approach

In a first approach, we compared the cumulative number of COVID-19 related deaths over the past 21 days relative to the capacity measures described in the previous paragraph. Yanget al. reported a median of 7 days in ICU for non-survivors, and at least 4 weeks for survivors [6]. Cumulative deaths over a 21 day time window therefore provide an approximation for the evolving burden of critical disease that is available in real time across EU countries, and which is likely less susceptible to inter-country variation in testing practices (and associated variations in underreporting).

We refer to the use of cumulative deaths over the last 21 days relative to health care system capacity as the “intensity-approach”. In a secondary approach, we compare the number of active COVID-19 cases, relative to the capacity measures. This latter approach is referred to as the “magnitude-approach”. Since the composite measure combines both health care staff and bed capacity, we focus our interpretation of pressure at the current stage of the epidemic in the first place on the intensity approach using the composite measure.

## Italy as a benchmark for healthcare pressure

We chose to set the situation in Italy on March 11, 2020 as a benchmark, based on Remuzzi & Remuzzi [1] and media reports [7]. This benchmark acts as a reference for a “soon to be overloaded” healthcare system.

We used the number of active COVID-19 cases and cumulative number of COVID-19 related deaths in Italy on March 11, 2020 to parameterize our benchmark. These numbers, 10 590 active cases and 827 deaths were derived from official Italian reporting data [8].

We retrieve the number of active reported cases and the cumulative number of COVID-19 related deaths over the past 21 days in real-time [9] and calculate the proportion per hospital bed, physician, the composite measures, and healthcare expenditures, respectively. We calculated, for each country, 16 ratios by dividing the number of cumulative deaths, and active reported cases, by each measure for both approaches. Afterwards, we normalized ratios by capacity measure benchmarks for Italy, which is defined as the cumulative number of COVID-19 related deaths (intensity-approach) or active cases (magnitude-approach) on March 11, divided by capacity measures that were derived for Italy. As such, when a country scores 1 on a specific measure, this reads as: “currently similar pressure as in Italy on March 11” for this specific measure. Since the results of these measures are very time-sensitive, we constructed a website that tracks healthcare pressure in real-time: www.covid-hcpressure.org

## Healthcare pressure indications in Europe derived on March 25, 2020

In Italy, the concentration of critical care beds is at the higher end with 12.5 critical care beds per 100 000 population, compared to the European average of 11.5 [5]. Moreover, the physician density was found to be above average, at about 400 physicians per 100 000 individuals [4]. In Figure 1, we displayed the results of the composite measures using the intensity-approach on March 25. We find that the pressure on the Italian healthcare system was 8 times higher on March 25 than on March 11. Spain is, next to Italy, the most severely affected country at about 7 times the pressure, compared to the benchmark situation. The Netherlands, France, Switzerland, The UK and Belgium’s health systems are also experiencing relatively high pressure, at similar or higher levels as in the benchmark situation. Note that these results can be consulted on the website and are updated every hour, for all combinations of measures and approaches.

**Figure 1:**
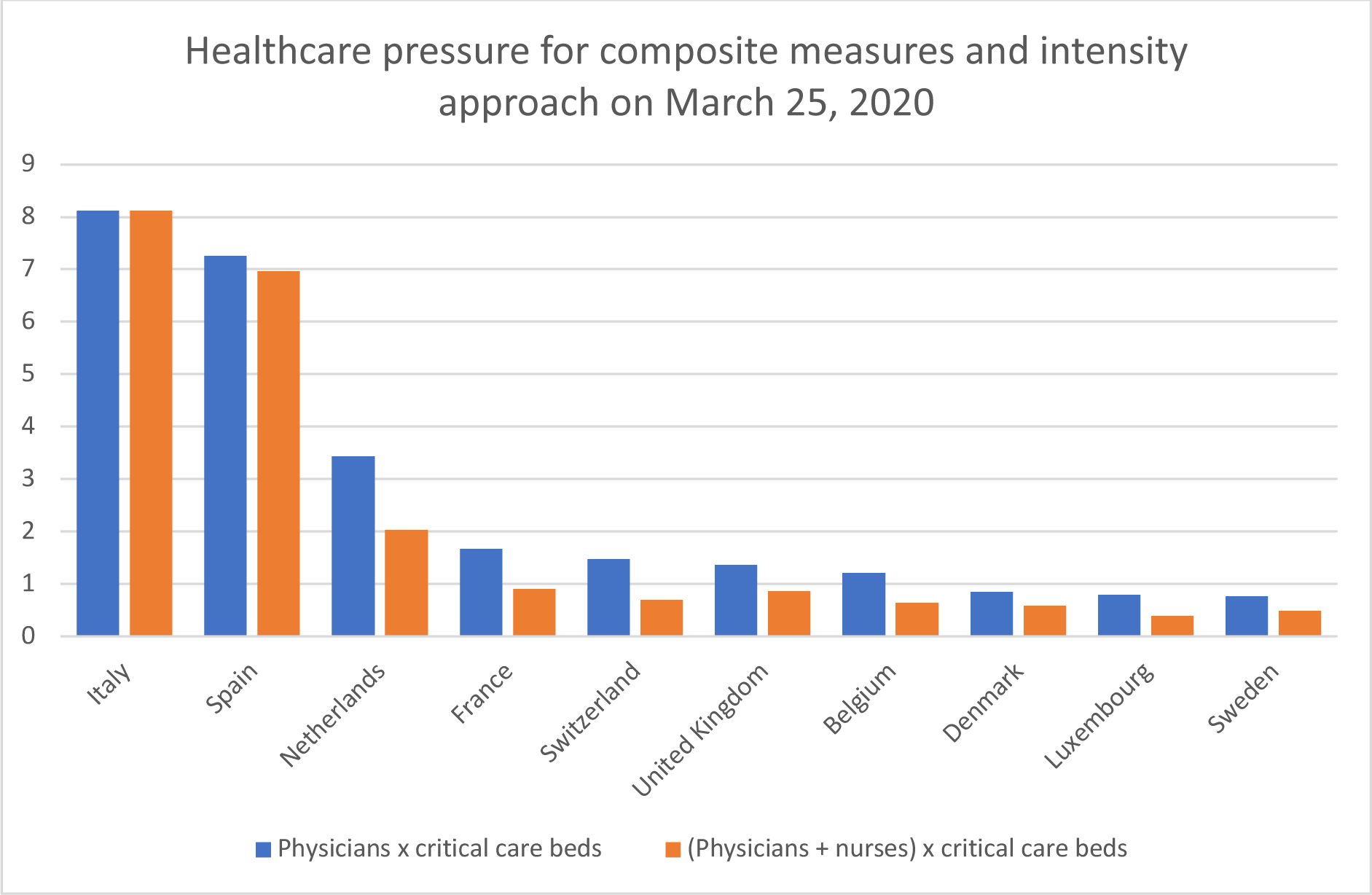
Relative pressure on the healthcare systems for 10 countries under relatively high pressure as indicated by composite measures, using the “intensity-approach”. Ratios are normalized to the cumulative number of COVID-19 related deaths in Italy on March 11, 2020. Ratios were calculated for the combination of critical care beds and either physicians, or physicians and nurses.

## How do countries compare to Italy?

The measures proposed in this paper are an approximation to the current healthcare capacity in Europe. Unfortunately, data on health care capacity is relatively scarce and we have to rely on data from 2017,2018 or even older. Data availability is even worse for critical care beds, on which we rely on multi-country data from 2010. Remuzzi & Remuzzi report ICU capacity of 5200 beds for Italy as a whole, which is lower than the 7550 critical care beds reported by Rhodes et al. [1,5]. It is likely that other EU countries have downsized their critical bed capacity too. In terms of curative beds per 100 000 population as reported by Eurostat, for most countries (all except Ireland, Bulgaria, Poland and Romania), we observed a downward trend in the past 10 years. Moreover, hospital capacity strain was recently found to be associated with increased mortality and decreased health outcomes [10]. Note that if we would assume the same extent of downsizing in Italy and other EU countries, our relative comparisons would not change. Note that we compare these countries by their base capacity, while clearly many countries have expanded their base capacity to differing degrees (eg, additional beds, mobilizing volunteers or retired health care workers). We therefore will track at which relative pressure individual countries can no langer handle the volume of critical patients and which pressure on their base capacity they experience at the peak. Note also that we do not account for health care workers’ incapacitation from COVID-19 or other causes as the epidemic unfolds. Our analysis shows that many European countries are soon to be confronted with a health care pressure that will exceed current healthcare capacity. Based on the intensity-approach and the composite measures, we believe that the healthcare pressure in Spain is already at very high levels. For The Netherlands and France the pressure on healthcare systems will soon reach Italy’s levels. Where this is not yet done, policymakers should urgently expand their health care capacities to avoid pressure as experienced in Italy. Our analysis informs policy makers on how far their country is removed from an overloaded healthcare system. As such, this information can be useful to plan ahead in order to relieve pressure on their national healthcare system. Moreover, the evolving healthcare pressure, especially the pressure experienced by each country at the peak, will be useful to plan future outbreaks and a timely expansion of health care capacity.

## Data Availability

The data used in the article is publicly available and referred to in the main text.

http://www.covid-hcpressure.org/home/

